# Causality of Abdominal Obesity on Cognition: a Trans-ethnic Mendelian Randomization study

**DOI:** 10.1101/2021.10.13.21264895

**Authors:** Shi-Heng Wang, Mei-Hsin Su, Chia-Yen Chen, Yen-Feng Lin, Yen-Chen A. Feng, Po-Chang Hsiao, Yi-Jiun Pan, Chi-Shin Wu

## Abstract

Obesity has been associated with cognition in observational studies; however, whether its effect is confounding, reverse causality, or causal remains inconclusive. Using two-sample Mendelian randomization (MR) analyses, we investigated the causality of overall obesity, measured by BMI, and abdominal adiposity, measured by waist–hip ratio adjusted for BMI (WHRadjBMI), on cognition. Using summary data from the GIANT consortium, COGENT consortium, and UK Biobank of European ancestry, there was no causal effect of BMI on cognition performance (beta[95% CI]=-0.04[-0.12,0.04], p-value=0.35); however, a 1-SD increase in WHRadjBMI was associated with 0.07 standardized decrease in cognition performance (beta[95% CI]=-0.07[-0.12,-0.02], p=0.006). Using raw data from the Taiwan Biobank of Asian ancestry, there was no causal effect of BMI on cognitive aging (beta[95% CI]=0.00[-0.09,0.09], p-value=0.95); however, a 1-SD increase in WHRadjBMI was associated with a 0.17 standardized decrease in cognitive aging (beta[95% CI]=-0.17[-0.30,-0.03], p=0.02). This trans-ethnic MR study reveals that abdominal adiposity impairs cognition.

## Introduction

Obesity is an important public health issue owing to its increasing prevalence ^1^ and adverse health effects on cardiovascular disease, diabetes, cancer, and depression ^2,3^. Mid-life obesity is a risk factor for the development of dementia in late life ^4^. Based on several observational studies, obesity is associated with reduced cognitive function in middle-aged adults ^5-7^. However, some studies failed to replicate such associations ^8,9^. Such an association may also be reverse causality. Childhood intelligence may be inversely associated with adult obesity ^10,11^. Whether the association of obesity with cognition is confounded by unmeasured factors or reverse causality remains inconclusive ^12,13^.

Mendelian randomization (MR) adopts the concept of a randomized control trial, and genetic variants can be used as instrumental variables for exposure of interest to uncover an observational association between an exposure and an outcome that is causal or simply correlated ^14^. The MR approach is not biased by reverse causation or confounding factors. Several studies have applied MR to demonstrate the effect of obesity on various physical complications, such as cardiovascular disease ^15,16^, cancer ^17^, and multiple sclerosis ^18^. Furthermore, studies have demonstrated a causal association between obesity and low gray matter volume ^19,20^. However, the causal association between obesity and cognition has not been examined using the MR approach.

The present study aimed to determine whether cognition is causally affected by genetically predicted overall obesity, measured by body mass index (BMI), and abdominal adiposity, measured by waist–hip ratio adjusted for BMI (WHRadjBMI), across populations, including European and Asian populations. We reported MR in a two-sample approach using genome-wide association study (GWAS) summary data for individuals of European ancestry, including BMI (n=322,154) and WHRadjBMI (n=210,088) from the GIANT consortium, and cognition performance (n=257,828) from the UK Biobank and COGENT consortium, and using individual data of Asian ancestry from Taiwan Biobank to perform GWAS for BMI (n=65,689), WHRadjBMI (n=65,683), and cognitive aging as measured by the Mini-Mental State Examination (MMSE, n=21,273).

## Results

### MR estimation of the causality of obesity on cognitive performance in Europeans

For the causality of BMI and WHRadjBMI on cognitive performance, a forest plot of the causal effect estimation for each instrument and the overall causal effect estimation by inverse variance weighted (IVW) is displayed in Figure 1 (a) and (b), respectively, and a scatter plot of MR is shown in Figure 1 (c) and (d), respectively. According to the main results using IVW, there was no causal effect of BMI on cognition (beta [95% CI] = -0.04 [-0.12, 0.04], p-value = 0.35, Figure 2); however, a 1-SD increase in WHRadjBMI was associated with 0.07 standardized score decrease in cognition (beta [95% CI] = -0.07 [-0.12, -0.02], p-value = 0.006).

**Figure 1.**
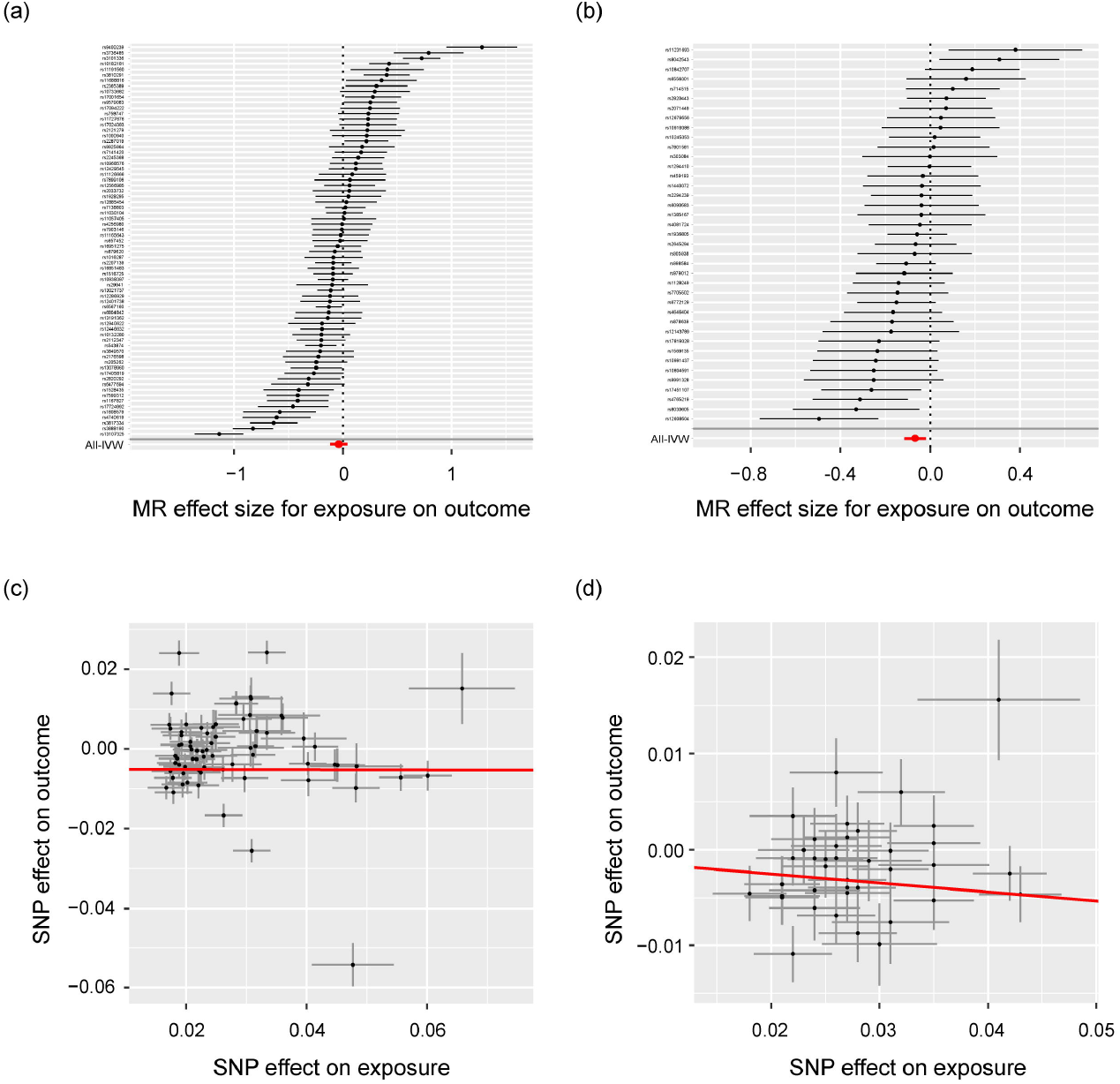
Forest plot of the causal effect estimation for a single instrument and the overall causal effect estimation by IVW, and a scatter plot of Mendelian randomization of obesity-related traits on cognitive performance in the European population. **(a)** forest plot for BMI; **(b)** forest plot for WHRadjBMI; **(c)** scatter plot for BMI; and **(d)** scatter plot for WHRadjBMI.

**Figure 2.**
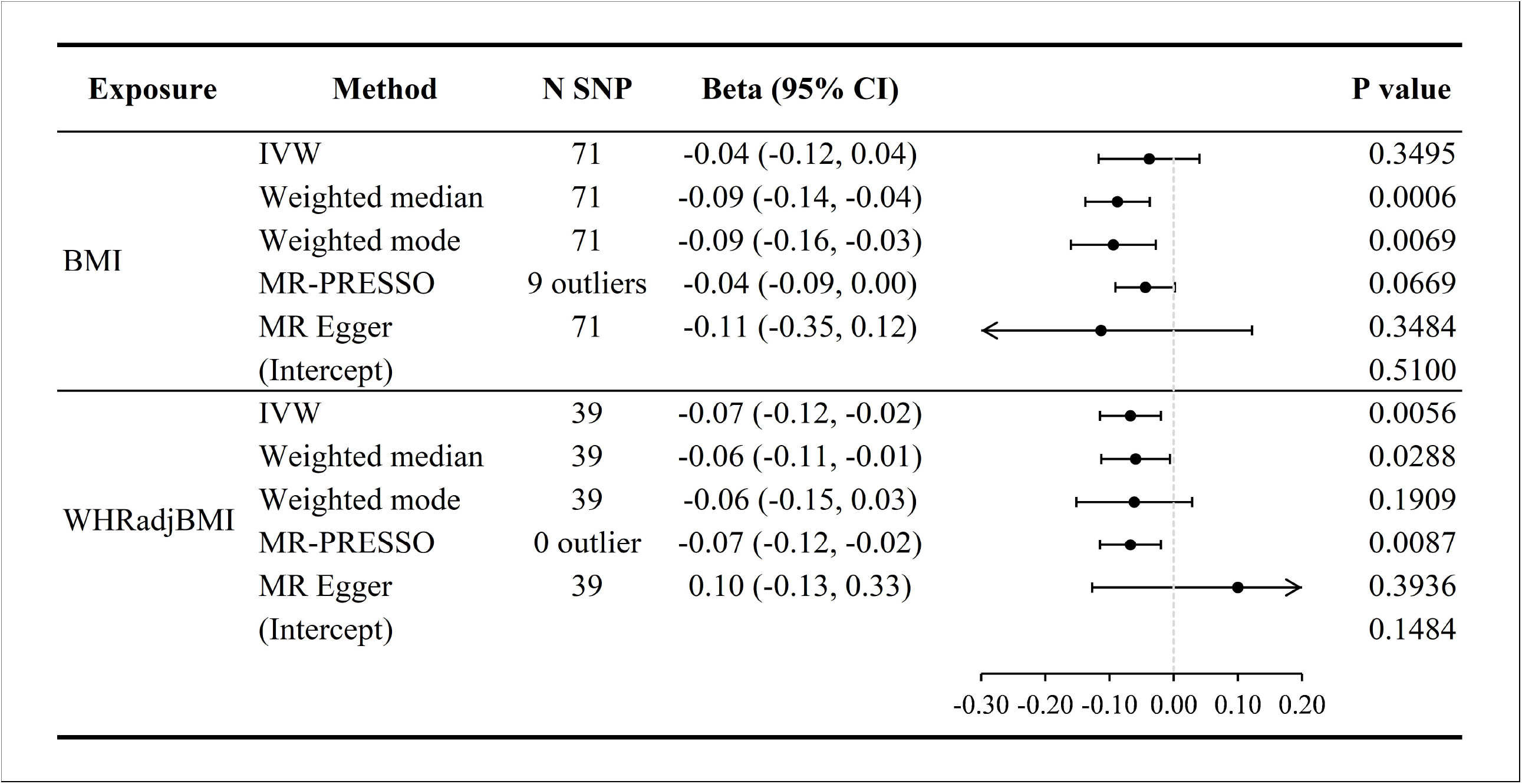
The causal effect estimates of obesity-related traits on cognitive performance in the European population using different Mendelian randomization methods.

Heterogeneity was detected across instrument effects in the causality of two obesity-related traits on cognitive performance (Supplementary Table S1), and overall pleiotropy was not detected using the MR-Egger intercept (Figure 2).

In the sensitivity analysis using other MR methods for BMI, the weighted median and weighted mode methods suggested that higher BMI was casually associated with lower cognition (p = 0.0006 and 0.007, respectively). However, after removing nine outliers, MR-PRESSO suggested no causality of BMI on cognition (p = 0.07).

In the sensitivity analysis for WHRadjBMI, the weighted median estimate suggested causality of WHRadjBMI on cognition (p = 0.03); however, MR-PRESSO did not detect any outliers and replicated the causality of WHRadjBMI on cognition (p = 0.0087).

### MR estimation of the causality of obesity on cognitive aging in Asians

For the causality of BMI and WHRadjBMI on cognitive aging, measured by MMSE, a forest plot and a scatter plot of MR are shown in Figure 3. Based on the main results using IVW, there was no causal effect of BMI on MMSE (beta [95% CI] = 0.00 [-0.09, 0.09], p-value = 0.95, Figure 4); however, a 1-SD increase in WHRadjBMI was associated with a 0.17 standardized score decrease in MMSE (beta [95% CI] = -0.17 [-0.30, -0.03], p-value=0.02).

**Figure 3.**
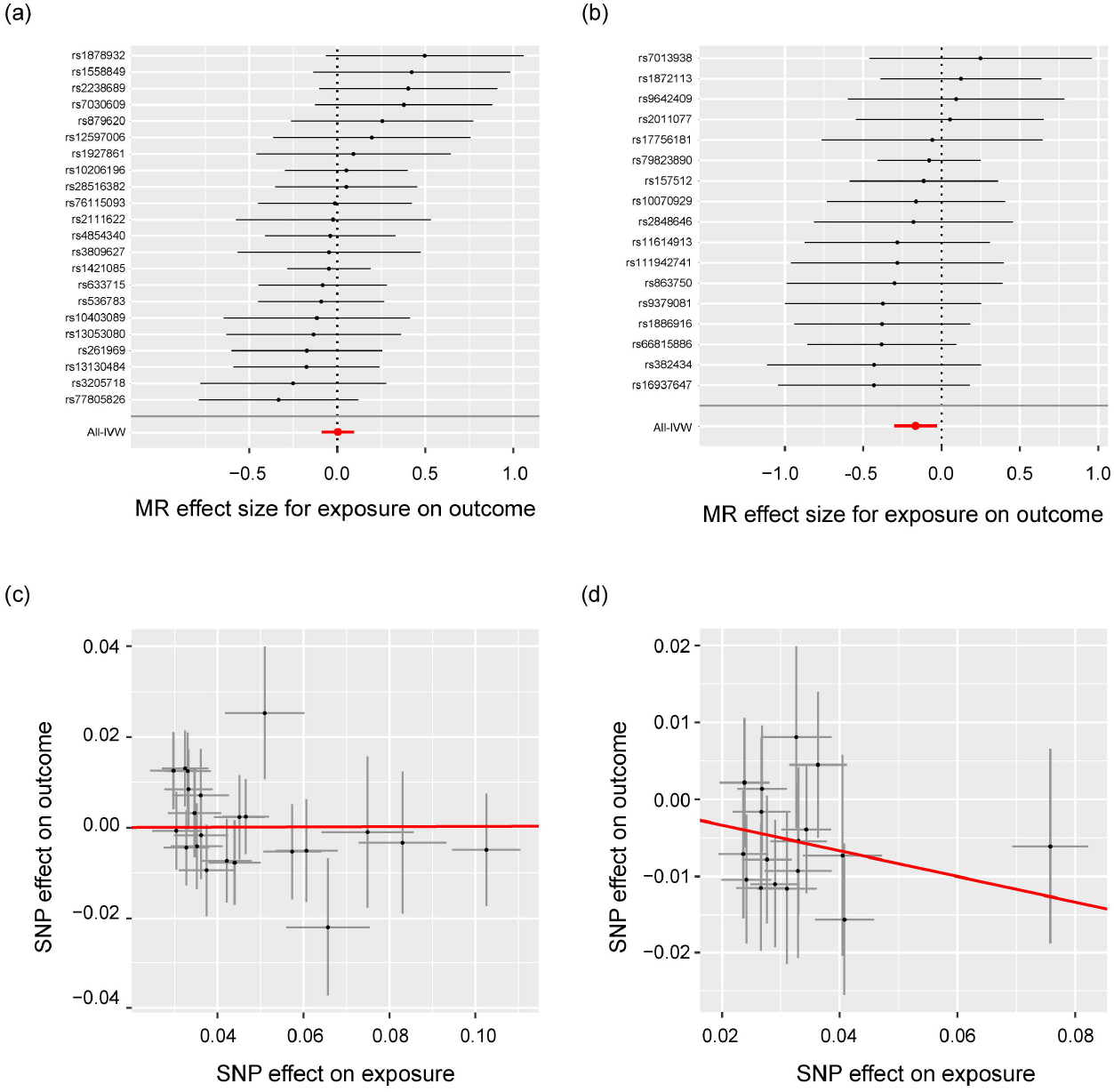
Forest plot of the causal effect estimation for a single instrument and the overall causal effect estimation by IVW, and a scatter plot of Mendelian randomization of obesity-related traits on MMSE in the Asian population. **(a)** forest plot for BMI; **(b)** forest plot for WHRadjBMI; **(c)** scatter plot for BMI; and **(d)** scatter plot for WHRadjBMI.

**Figure 4.**
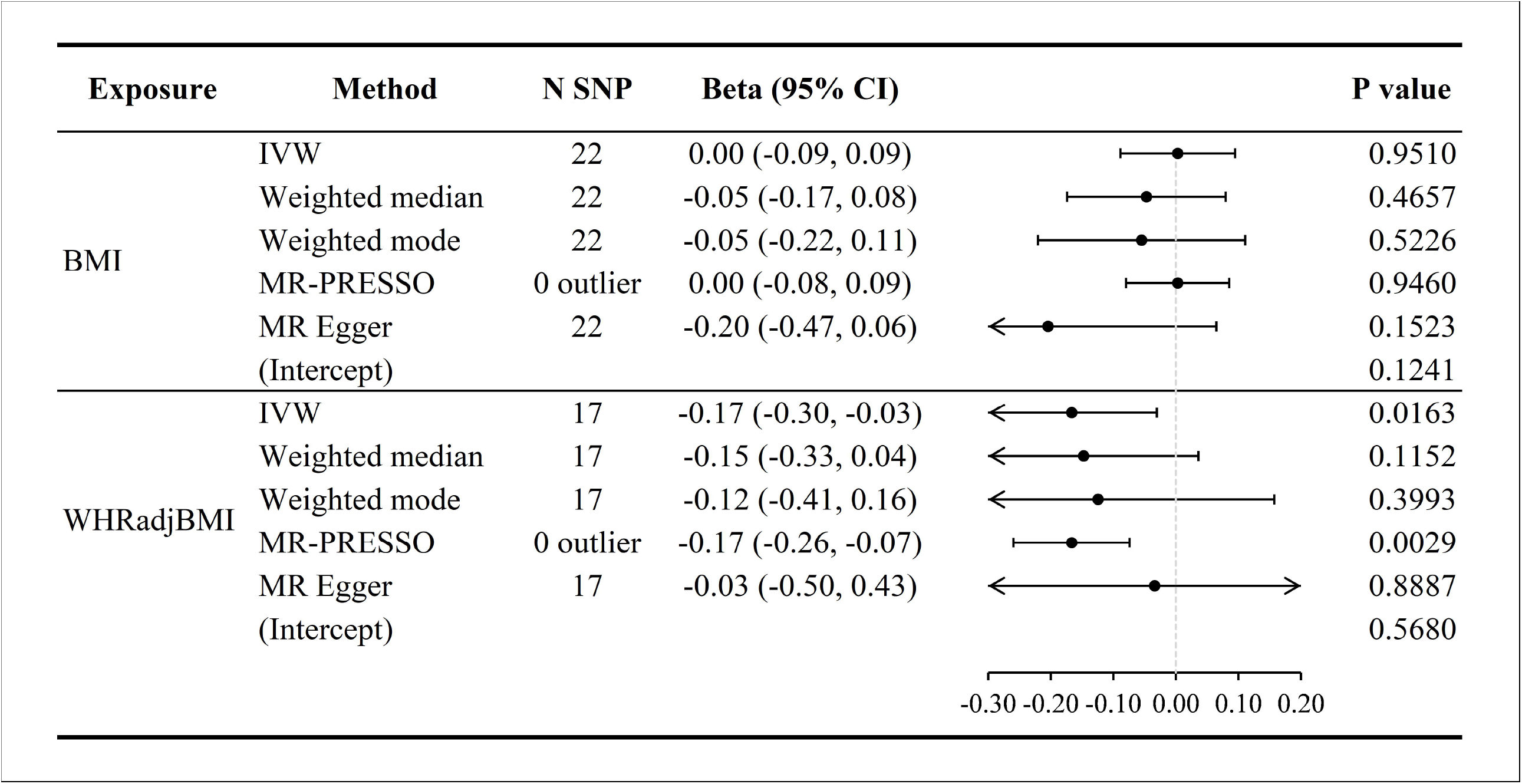
The causal effect estimates of obesity-related traits on MMSE in the Asian population using different Mendelian randomization methods.

For the effect of both BMI and WHRadjBMI on cognitive aging, heterogeneity was not detected across the instrument effect (Supplementary Table S3), and overall pleiotropy was not detected using the MR-Egger intercept (Figure 4).

In the sensitivity analysis for BMI, none of the MR estimates supported the causality of BMI on cognitive aging. In the sensitivity analysis for WHRadjBMI, MR-PRESSO did not detect any outliers and replicated the causality of WHRadjBMI on MMSE (p = 0.003). The weighted median and weighted modes had similar causality estimations, although no statistical significance was found.

## Discussion

Our study confirmed a causal relationship, as abdominal adiposity measured by WHRadjBMI was found to have an adverse effect on cognition across European and Asian populations. Each SD increase in genetically predicted WHRadjBMI was associated with greater than 7% decrease in the standardized performance of cognition in Europe and a 17% decrease in the standardized performance of cognitive aging in Asians. However, weak evidence is provided for the causal relationship between BMI and cognition in both populations.

Abdominal adiposity, measured by WHRadjBMI, was found to be associated with poor cognition. Our findings are consistent with those of previous observational studies ^21-24^. One cross-sectional study assessed the effect of both BMI and WHR on cognition in older adults. This study revealed that only individuals with BMI >28 kg·m^-2^ and high WHR 0.8 had statistically significant cognitive impairment ^21^. Another study revealed no association between BMI and cognition among Chinese elderly individuals; however, interactions were found between BMI and WHR on cognition performance. In fact, the researchers found that high WHR was associated with cognitive impairment only among those with BMI > 25.3 kg·m^-2 24^. Another study used both BMI and WHR as obesity indicators and found that the upper quartile of WHR in midlife predicted poor cognitive function 12 years later. However, there was no significant difference between BMI and < 30 kg·m^-2 22^. The Women’s Health Initiative Memory Study found that women with high WHR and normal BMI had a higher risk of developing cognitive impairment than those with BMI 35 kg·m^-2 23^. In addition to cognition, several studies have demonstrated that WHR might be a better tool for assessing cardiovascular disease and other health outcomes ^25,26^.

In the present study, no association was found between BMI and cognition. Our findings were consistent with those of previous studies that failed to demonstrate such an association in the full adjustment model. ^8,9,22^. However, the association between BMI and cognition is inconsistent in the literature. Several studies have revealed an association between baseline obesity and poor cognitive performance at follow up ^5,27,28^. BMI is composed of fat, water, muscle, bone, and other tissues, which may have different effects on health outcomes. Therefore, BMI might be a relatively poor biological tool for the examination of causal pathways in diseases ^29^.

Several MR studies have revealed that obesity causes a low volume of gray matter, which indicates that obesity is associated with neurodegeneration ^19,20^. However, some MR studies did not reveal the effect of BMI on the incidence of Alzheimer’s disease ^30-32^. Among these studies, two used BMI as an obesity indicator only ^30,31^. Only one study included BMI, WHRadjBMI, and waist circumference. Although none of these indicators were significantly associated with Alzheimer’s disease, the strength of association for WHRadjBMI was higher than that for other indicators ^32^. We hypothesized that the etiology of Alzheimer’s disease is mainly caused by the abnormal build-up of neurofibrillary tangles and beta-amyloid plaques ^33^. However, the adverse effect of obesity on cognition might be due to the cardiometabolic system, including cerebrovascular diseases. Obesity decreases not only blood supply to the brain, but also increases fat cells that damage the brain white matter, leading to loss of cognitive and intellectual behavior ^34^. Of note, a meta-analysis of clinical trials demonstrated that intentional weight loss might be associated with improvement of executive function and memory in obese individuals ^35^. However, the sample size of all trials was small. Accordingly, the effectiveness of body weight reduction in enhancing cognition should be replicated in a large-scale study.

The strengths of this study include the use of the MR approach to overcome confounding and reverse causality in conventional observational studies, and the use of large-scale GWAS to achieve a well-powered MR analysis, and trans-ethnic analysis to improve generalizability. Nonetheless, this study had several limitations. First, similar to all MR studies, the potential influence of pleiotropy on the results cannot be ruled out entirely. However, the genetic association between obesity and cognition was similar in the sensitivity analyses, and there was no evidence of pleiotropy. Second, the effect of modification on the link between obesity and cognition was not considered in this study. Thus, we could not assess the association between adiposity and cognition for different age groups and genders, with potentially different fat accumulation in different locations. Third, the measures for cognition tests differed across the populations in our analyses. Only one representative indicator was included for the given population in our analysis. The effect of obesity on specific executive functions could not be explored in detail.

In conclusion, the findings from this trans-ethnic MR study indicate that genetically predicted abdominal adiposity, as measured by WHR adjusted for BMI, would impair cognition. However, the mechanism underlying causality warrants further investigation. Healthcare providers may recommend avoiding abdominal obesity to prevent cognitive deficits and resist cognitive aging. Further investigations should focus on whether intentional weight reduction can reverse the adverse effects on cognition.

## Methods

### Data Source for Europeans

This study used publicly available GWAS summary meta-analysis statistics for obesity-related traits from the GIANT consortium and cognition performance from the UK Biobank and COGENT consortium for individuals of European ancestry.

#### GWAS for exposure: obesity traits

BMI is the most commonly used measure of overall obesity. However, BMI does not reflect the proportion of weight related to muscle or fat within the body. The variation in body fat distribution could be high for a given BMI. WHRadjBMI is a surrogate measure of abdominal adiposity ^36^. A GWAS summary for obesity-related phenotypes, including BMI and WHRadjBMI, was retrieved from a meta-analysis of the GIANT consortium with approximately 25 million genetic variants. A GWAS for BMI was conducted among 322,154 individuals of European descent ^37^. GWAS for WHRadjBMI was conducted among 210,088 individuals of European descent ^38^. The obesity-related variables underwent a standardized transformation by the inverse standard normal function before the association tests.

#### GWAS for outcome: cognitive performance

The GWAS summary statistics for cognition performance were retrieved from a meta-analysis of the UK Biobank and COGENT consortium ^39^, with a total of 257,828 individuals. The UK Biobank used a standardized score on a verbal-numerical reasoning test. The test contains 13 logic and reasoning questions with a two-minute time limit and was designed as a measure of fluid intelligence, which indicates the ability to think and reason abstractly and solve problems. Each participant took the test four times. The mean scores were used and standardized. The COGENT consortium defined cognition phenotype as the first unrotated principal component of performing multiple neuropsychological tests, including IQ-test subscales, across 35 studies. In general, the test variables measured the overall accuracy or total number of correct responses. The average Cronbach’s alpha (a measure of internal consistency between test items) was 0.70 across component studies.

### Data Source for Asians

This study used individual genotyping and phenotyping data from the Taiwan Biobank, the largest government-supported biobank in Taiwan since 2012 ^40^. The Taiwan Biobank collects community-based samples for individuals between 30 and 70 years who are cancer-free at recruitment. The recruitment and data collection procedures were approved by the internal review board of the Taiwan Biobank. Each subject signed an approved informed consent form and provided blood samples, underwent physical examinations, and participated in face-to-face interviews. This study was approved by the Central Regional Research Ethics Committee of China Medical University, Taichung, Taiwan (CRREC-108-30).

This study comprised 95,238 individuals, including 34,595 males and 60,643 females, for whom genome-wide genotyping was carried out using the custom Taiwan Biobank chips ran on the Axiom Genome-Wide Array Plate System (Affymetrix, Santa Clara, CA, USA); 26,274 participants were genotyped on the TWBv1 chip and 68,964 participants were genotyped on the TWBv2 chip. Quality control and imputation of the two chips were conducted separately. Quality control included the exclusion of individuals with more than 5% missing variants, exclusion of variants with a call rate <5%, minor allele frequency (MAF) <0.001, and deviation from Hardy-Weinberg equilibrium with P <1 × 10^−6^. We used the 504 EAS panel from 1000 Genomes Project ^41^ and the 973 TWB panel from whole-genome sequencing in TWB participants as the reference panel to impute the genotypes with IMPUTE2 and then retained the variants with imputation info > 0.7 and MAF > 0.5%. We removed multi-allelic variants and variants located in long-range LD regions (chr6: 25-35Mb; chr8: 7-13Mb).

Two independent sample sets were selected for GWAS for cognition and GWAS for obesity, one with samples ≥ 60y having information on cognitive aging test and the other with samples < 60y having information on obesity-related traits. In each sample set, cryptic relatedness was removed, and the identity by descent (IBD) sharing coefficients, PI-HAT = probability (IBD = 2) + 0.5× probability (IBD = 1), between any two participants in KING, and excluded one individual from a pair with PI-HAT greater than 0.1875. We performed principal component analyses (PCA) to identify population outliers; samples with any of the top 20 principal components (PCs) more than 6 SD away from the sample average were removed.

For each trait, we removed samples with measures that were more than 6 SD away from the sample mean. We then normalized each trait by performing an inverse rank-based normal transformation. We performed a genetic association test separately for the two chips, and then performed an inverse-variance-weighted fixed-effect meta-analysis in METAL. For each trait, the sample size and heritability estimates for each chip, and genetic correlation estimates between the two chips are provided in Supplementary Table S2.

#### GWAS for exposure: obesity traits

For BMI with a sample size of 65,689, we performed linear regression in PLINK for association tests with adjustment for age, age^2^, sex, age by sex interaction, age^2^ by sex interaction, and top 20 PCs. We performed an association test for WHRadjBMI with a sample size of 65,683. Manhattan plots for BMI and WHRadjBMI are shown in Supplementary Figure 1.

#### GWAS for outcome: cognitive aging

The Mini-Mental State Examination (MMSE)^42^, the most commonly used tool for testing cognitive aging, was measured using a questionnaire employed during face-to-face interviews with subjects > 60 years. The MMSE included tests for orientation, memory attention, calculation, and language function. We performed an association test for MMSE with adjustment for age, age^2^, sex, age by sex interaction, age^2^ by sex interaction, top 20 PCs, and educational attainment with a sample size of 21,273. The Manhattan plot for MMSE is shown in Supplementary Figure 1.

### Selection of genetic variants for MR

Genetic instruments for the exposure of interest were selected based on the same processes for each obesity-related phenotype. We first mapped the variants from the exposure data to the outcome GWAS and preserved those that could be mapped to both. To ensure that the extracted variants were independent, linkage disequilibrium (LD) clumping was conducted based on r^2^ > 0.0001 within a 1000 kb window. Variants with genome-wide significance (p < 5×10^−8^) for exposure were selected as genetic instruments. After harmonization and removing palindromic variants, the available instrumental variables were 71 for BMI and 39 for WHRadjBMI in the European data, and 22 for BMI and 17 for WHRadjBMI in the Asian data. For Asians, information on the association of individual genetic instrument with obesity traits and with MMSE estimated from the Taiwan Biobank is provided in Supplementary Table S3.

### MR approach for causality estimation

We performed MR analysis using the IVW method for the main results ^43^ as it is the best unbiased estimation if there is no pleiotropy and all instruments are assumed to be valid ^44^. Several different MR methods were used for the sensitivity analysis. Pleiotropy indicates that one gene manifests two or more unrelated phenotypes. The association between the gene and the outcome might occur through different pathways rather than the phenotype of interest. Therefore, we employed MR-Egger ^44^, weighted median ^45^, and weighted mode ^46^ methods to achieve robust and more reliable estimations while pleiotropy existed. The MR-Egger regression considers pleiotropy. The weighted median provides a consistent estimation even when 50% of the information is derived from invalid instrumental variables. The weighted mode is consistent even if most of the instruments are invalid. MR PRESSO was also conducted to detect and adjust for possible outliers ^47^. R version 3.6.0 and TwoSampleMR version 0.5.3 for Linux were used for the MR analyses.

We assessed the heterogeneity between individual instrument-cognition on instrument-obesity using Cochran’s and Rucker’s Q test ^48,49^, and examined the overall pleiotropy for each MR analysis using the MR-Egger intercept.

## Data Availability

All data produced are available online at UK Biobank & Taiwan Biobank.

https://www.twbiobank.org.tw/

https://www.ukbiobank.ac.uk/

## Acknowledgements

This work was supported by National Health Research Institutes (NHRI-EX109-10931PI, NHRI-EX110-10931PI).

## Author contributions

Conceptualized the study: SHW, CSW.

Performed the analysis: SHW, MHS, PCH.

Wrote the paper: SHW, CSW.

Critically revised the paper: MHS, CYC, YFL, YAF, PCH, YJP.

All authors reviewed and approved the final version of the manuscript.

## Competing interests

Chia-Yen Chen is an employee of Biogen. The remaining authors declare no conflict of interest.

## Notes

### Competing Interest Statement

The authors have declared no competing interest.

### Author Declarations

This study was approved by the Central Regional Research Ethics Committee of China Medical University, Taichung, Taiwan (CRREC-108-30).

