## Supplementary Table S1∼S3 & Figure S1 for "Causality of Abdominal Obesity on Cognition: a Trans-ethnic Mendelian Randomization study"

Supplementary Table S1. Heterogeneity test across instrument effect of obesity-related traits on cognition.

| Population | Exposure | IVW | |  | MR-Egger | |
| --- | --- | --- | --- | --- | --- | --- |
|  |  | Cochran’s Q | p |  | Rücker's Q | p |
| European | BMI | 555.89 | <0.0001 |  | 555.37 | <0.0001 |
| European | WHRadjBMI | 67.99 | 0.0020 |  | 64.20 | 0.0036 |
| Asian | BMI | 14.3174 | 0.8140 |  | 16.8946 | 0.7175 |
| Asian | WHRadjBMI | 7.1342 | 0.9538 |  | 7.4751 | 0.9630 |

Supplementary Table S2. Sample size and heritability estimates within batch 1 (TWBv1 chip) and batch 2 (TWBv2 chip), and genetic correlation estimates between the two batches of the Taiwan Biobank samples in body mass index (BMI), waist–hip ratio adjusted for BMI (WHRadjBMI), and Mini-Mental State Examination (MMSE).

|  | TWBv1 chip | | | TWBv2 chip | | | Genetic Correlation | | |
| --- | --- | --- | --- | --- | --- | --- | --- | --- | --- |
| Trait | Sample size | h*^2^_g_* (SE) | λ_GC_ | Sample size | h*^2^_g_* (SE) | λ_GC_ | r_g_ | SE | P-value |
| BMI | 18579 | 0.2162 (0.0295) | 1.0895 | 47110 | 0.2318 (0.0153) | 1.2038 | 1.0101 | 0.0774 | 6.2683E-39 |
| WHRadjBMI | 18578 | 0.1011 (0.0264) | 1.0375 | 47105 | 0.1181 (0.0146) | 1.1113 | 1.2788 | 0.186 | 6.17E-12 |
| MMSE | 5343 | 0.0523 (0.0852) | 1.0135 | 15930 | -0.0011 (0.0274) | 1.0016 | NA | NA | NA |

Supplementary Table S3. Association of individual SNPs with obesity traits and with MMSE in Taiwan Biobank.

| Exposure | Outcome | SNP | effect_allele | other_allele | beta_exposure | beta_outcome | pval_exposure | pval_outcome |
| --- | --- | --- | --- | --- | --- | --- | --- | --- |
| BMI | MMSE | rs1421085 | T | C | -0.1025 | 0.0049 | 8.51E-39 | 0.6917 |
| BMI | MMSE | rs10206196 | T | C | -0.0466 | -0.0024 | 3.08E-18 | 0.7696 |
| BMI | MMSE | rs536783 | T | C | 0.0574 | -0.0053 | 5.30E-18 | 0.6133 |
| BMI | MMSE | rs633715 | T | C | -0.0607 | 0.0051 | 1.72E-17 | 0.6493 |
| BMI | MMSE | rs4854340 | A | G | 0.0830 | -0.0033 | 2.02E-16 | 0.8311 |
| BMI | MMSE | rs28516382 | T | C | 0.0451 | 0.0023 | 1.05E-14 | 0.8046 |
| BMI | MMSE | rs13130484 | T | C | 0.0440 | -0.0077 | 1.12E-13 | 0.4093 |
| BMI | MMSE | rs261969 | A | C | -0.0422 | 0.0073 | 1.72E-13 | 0.4291 |
| BMI | MMSE | rs76115093 | T | C | 0.0749 | -0.0010 | 1.28E-12 | 0.9499 |
| BMI | MMSE | rs77805826 | T | C | -0.0657 | 0.0220 | 6.30E-12 | 0.1486 |
| BMI | MMSE | rs13053080 | T | C | 0.0328 | -0.0044 | 3.03E-10 | 0.5951 |
| BMI | MMSE | rs7030609 | A | G | -0.0331 | -0.0125 | 4.40E-10 | 0.1408 |
| BMI | MMSE | rs2238689 | T | C | 0.0325 | 0.0131 | 8.80E-10 | 0.1187 |
| BMI | MMSE | rs879620 | T | C | 0.0333 | 0.0085 | 1.37E-09 | 0.3359 |
| BMI | MMSE | rs10403089 | T | C | -0.0352 | 0.0041 | 2.22E-09 | 0.6646 |
| BMI | MMSE | rs3205718 | T | C | 0.0375 | -0.0094 | 3.98E-09 | 0.3540 |
| BMI | MMSE | rs3809627 | A | C | -0.0362 | 0.0017 | 4.57E-09 | 0.8567 |
| BMI | MMSE | rs1927861 | T | C | 0.0347 | 0.0032 | 1.09E-08 | 0.7460 |
| BMI | MMSE | rs1878932 | A | G | 0.0510 | 0.0253 | 1.48E-08 | 0.0825 |
| BMI | MMSE | rs2111622 | A | G | 0.0304 | -0.0007 | 1.63E-08 | 0.9370 |
| BMI | MMSE | rs1558849 | T | G | 0.0298 | 0.0126 | 1.71E-08 | 0.1373 |
| BMI | MMSE | rs12597006 | T | C | -0.0361 | -0.0071 | 2.25E-08 | 0.4897 |
| WHRadjBMI | MMSE | rs79823890 | T | G | -0.0758 | 0.0061 | 1.00E-31 | 0.6317 |
| WHRadjBMI | MMSE | rs157512 | T | C | -0.0343 | 0.0039 | 1.59E-16 | 0.6365 |
| WHRadjBMI | MMSE | rs66815886 | T | G | -0.0408 | 0.0156 | 1.68E-16 | 0.1125 |
| WHRadjBMI | MMSE | rs1872113 | A | G | -0.0363 | -0.0045 | 6.40E-14 | 0.6369 |
| WHRadjBMI | MMSE | rs1886916 | T | C | 0.0290 | -0.0110 | 2.51E-12 | 0.1831 |
| WHRadjBMI | MMSE | rs10070929 | T | G | 0.0330 | -0.0054 | 3.10E-12 | 0.5689 |
| WHRadjBMI | MMSE | rs11614913 | T | C | 0.0276 | -0.0078 | 2.49E-11 | 0.3504 |
| WHRadjBMI | MMSE | rs2011077 | T | C | 0.0268 | 0.0014 | 8.80E-11 | 0.8655 |
| WHRadjBMI | MMSE | rs16937647 | T | G | 0.0266 | -0.0115 | 1.20E-10 | 0.1683 |
| WHRadjBMI | MMSE | rs9379081 | T | G | 0.0310 | -0.0116 | 6.07E-10 | 0.2417 |
| WHRadjBMI | MMSE | rs2848646 | T | C | 0.0405 | -0.0073 | 7.48E-10 | 0.5773 |
| WHRadjBMI | MMSE | rs111942741 | T | C | -0.0329 | 0.0093 | 3.92E-09 | 0.4121 |
| WHRadjBMI | MMSE | rs382434 | T | C | -0.0241 | 0.0104 | 4.79E-09 | 0.2151 |
| WHRadjBMI | MMSE | rs9642409 | T | C | -0.0238 | -0.0022 | 7.28E-09 | 0.7933 |
| WHRadjBMI | MMSE | rs863750 | T | C | 0.0236 | -0.0071 | 9.60E-09 | 0.3913 |
| WHRadjBMI | MMSE | rs17756181 | A | G | 0.0267 | -0.0016 | 2.53E-08 | 0.8717 |
| WHRadjBMI | MMSE | rs7013938 | A | C | 0.0326 | 0.0081 | 2.77E-08 | 0.4908 |

Supplementary Figure 1. Manhattan plots of association test for (a) body mass index (BMI), (b) waist–hip ratio adjusted for BMI (WHRadjBMI), and (c) Mini-Mental State Examination (MMSE) in Taiwan Biobank.

| (a)  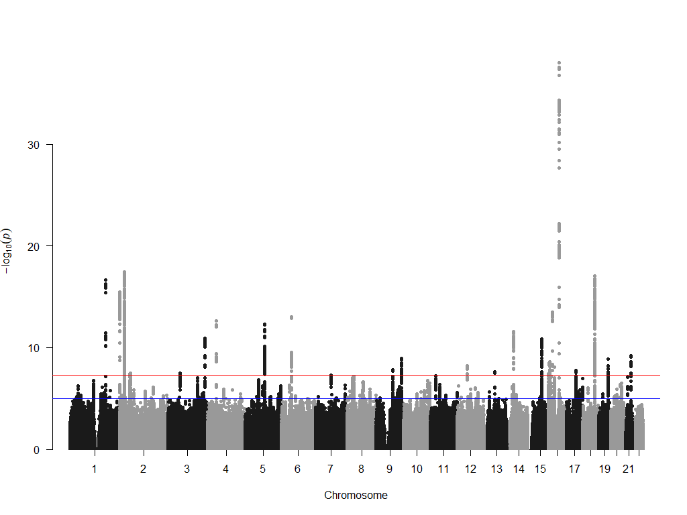 | (b)  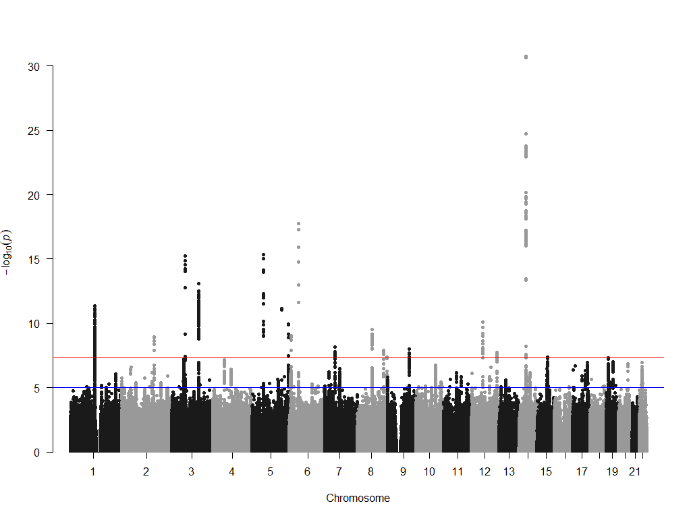 | (c)  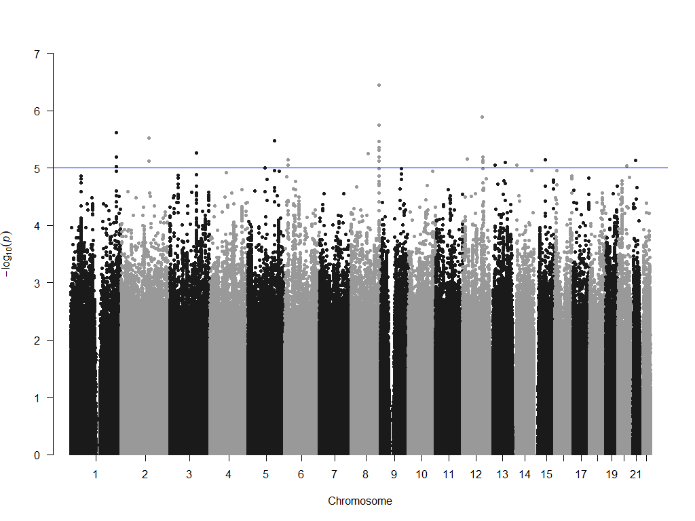 |
| --- | --- | --- |
